# Evaluation of EUROSCORE II to determine the prognosis of patients with moderate-to-severe aortic stenosis: a long-term retrospective study

**DOI:** 10.1101/2024.08.23.24312516

**Authors:** Xianzhen Cai, Danchun Hu, Baoxin Yan, Jinhao Chen, Weiwen Li, Ying Lin, Run Lin, Junjun Ye, Xiaojun Huang, Bin Xie, Xiaodong Zhuang, Jianying Chen, Jilin Li

## Abstract

**Background and Aims:** Aortic stenosis (AS) was a prevalent heart valve disease whose morbidity and mortality can be reduced by aortic valve replacement (AVR) . EUROSCORE II assesses the perioperative mortality of severe AS patients undergoing AVR. This study explored EUROSCORE II’s prognostic value for long-term all-cause mortality of Chinese patients with moderate-to-severe AS and determined whether AVR affects this.

**Methods:** Allocated to four groups following the EUROSCORE II (cut-off value of 4%) and whether performed AVR, 544 patients with moderate-to-severe AS were enrolled. Baseline data, Kaplan–Meier, Cox regression and subgroup analysis were used to analyse the relationship between EUROSCORE II and participants’ all-cause mortality. Furthermore, ROC analysis determining the optimal cut-off value of EUROSCORE II was utilized.

**Results:** During a median follow-up of 41.4 months, 177 (21.5%) participants reached the endpoint, with higher risks (EUROSCORE II ≥4%) and no AVR exhibited significantly increased all-cause mortality rates compared to other groups (55.4% vs. 6.5%, 13.4%, and 32.7%; P<0.001). Kaplan-Meier curves confirmed these findings (log-rank test P<0.001). Cox regression analysis revealed a 6.891-fold higher risk (HR, 6.891; 95% CI, 3.083-15.401; P<0.001) in patients without AVR with higher EUROSCORE II. The adjusted model (P<0.01) and subgroup analyses (without AVR P=0.001; with AVR P=0.029) supported EUROSCORE II’s prognostic value for all-cause mortality. The optimal EUROSCORE II cut-off for predicting all-cause mortality in patients without AVR was 2.23% (AUC 0.675).

**Conclusions:** EUROSCORE II (cut-off value 4%) and AVR independently impact the long-term prognosis of patients with moderate-to-severe AS.

## Introduction

Aortic stenosis (AS) is the most prevalent heart valve disease, causing significant morbidity and mortality in the elderly due to aging of the population[1-3]. Its prevalence exceeds 10% in individuals aged over 65 in US and European populations[4-6], although the prevalence in China may be lower, according to the results of a single-centre retrospective study of the echocardiographic data of 287,556 patients[7]. The initial stages of the disease are characterized by the progression of valvular lesions, involving endothelial cell damage, infiltration with lipids and macrophages, lipid oxidation, and subsequent fibrosis and calcification, which ultimately leads to obstruction of the aortic valve[6,8].

To date, owing to the absence of specific medication for the treatment or prevention of the progression of AS, surgical aortic valve replacement (SAVR) or transcatheter aortic valve implantation (TAVI) are the recommended means of treating patients with severe symptoms[9,10]. EUROSCORE II is a widely-used risk assessment tool[11-17] that uses various characteristics of the patient to estimate the risk of adverse outcomes, and it has been clinically implemented to identify high-risk patients during hospitalization and predict 30-day mortality in those undergoing coronary artery bypass grafting (CABG)[18], SAVR, or TAVR[19]. However, data are lacking regarding the long-time prognostic value of EUROSCORE II with respect to the mortality and morbidity rates of patients with moderate-to-severe AS, and whether AVR (SAVR or TAVR) is associated with a superior prognosis, especially in Chinese patients.

Consequently, the primary objective of this retrospective study was to characterize the relationships of EUROSCORE II and AVR status with all-cause mortality in patients with moderate-to-severe AS. In addition, we aimed to identify the optimal cut-off value of EUROSCORE II for prognostic use in patients who do not undergo AVR.

## Methods

### Study design and participants

A total of 1,033 patients diagnosed with moderate-to-severe AS at the Second Affiliated Hospital of Shantou University Medical College, the First Affiliated Hospital of Sun Yat-sen University, or the Affiliated Hospital of Guangdong Medical University were enrolled between January 2014 and July 2023. Three heart valve centers are derived from a database called Aortic Valve Diseases RISk facTOr assessmenT and Prognosis modeL Construction (ARISTOTLE). The primary outcome was all-cause mortality during the follow-up period. The inclusion criteria were as follows: (1) moderate-to-severe AS (peak aortic jet velocity (Vmax) >3 m/s, mean aortic pressure gradient (MG) >20 mmHg, or aortic valve area (AVA) ≤1.5 cm^2^) on echocardiography, according to the 2021 European Society of Cardiology (ESC)/ European Association for Cardio Thoracic Surgery (EACTS) guidelines for the management of valvular heart disease [9]; (2) congenital stenosis of the aortic valve; (3) an initial diagnosis of aortic stenosis; and (4) no previous history of AVR. The exclusion criteria were as follows: (1) a history of aortic valve replacement; (2) severe dysfunction or malignancy of other organs or systems; (3) connective tissue disease; (4) aortic stenosis in combination with severe mitral stenosis; (5) malignant tumour; and (6) missing baseline data (N = 419). Ultimately, 544 patients with moderate-to-severe AS remained for the analysis (Fig. 1).

**Fig. 1.**
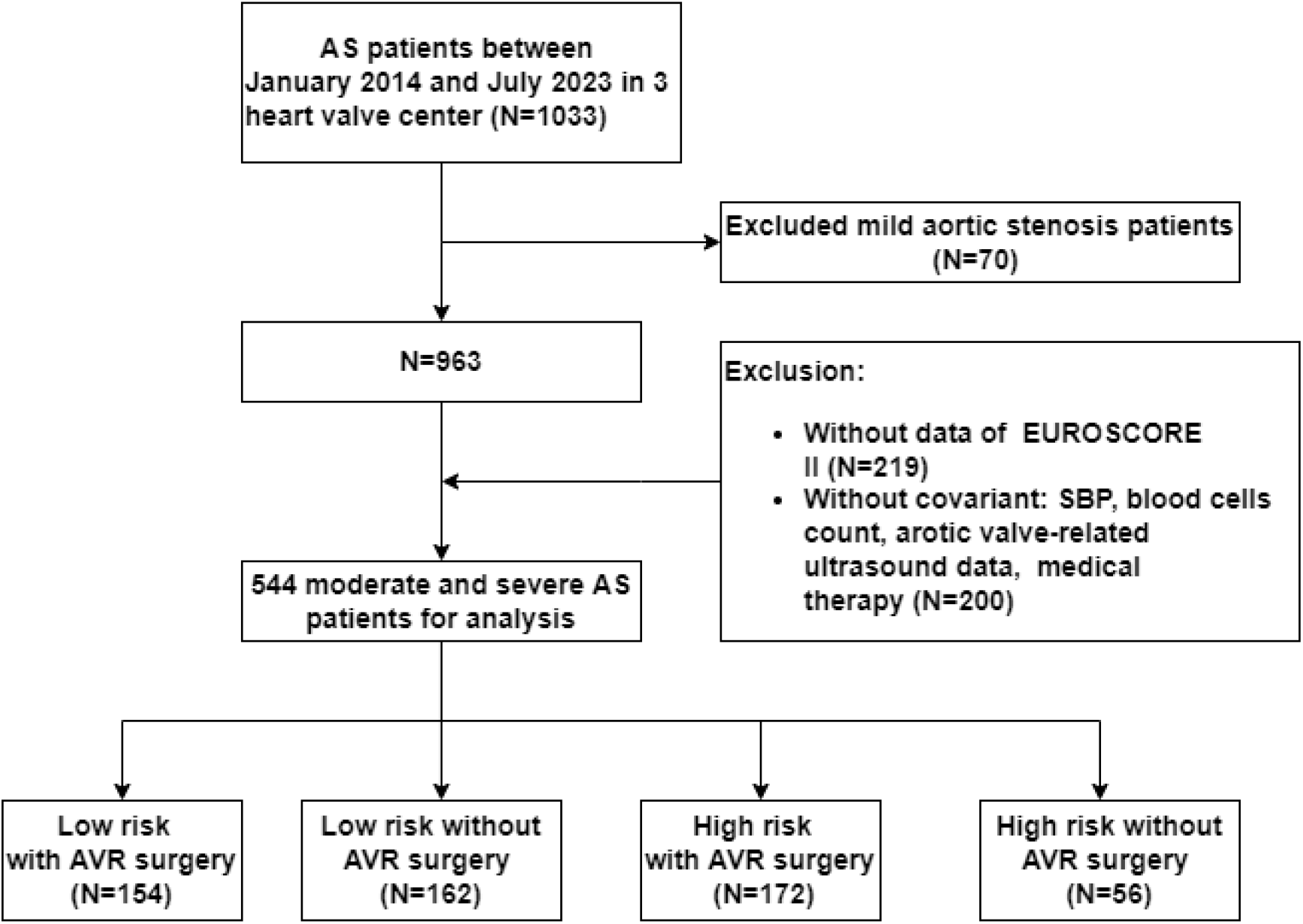
Flow diagram describing the study sample The study was performed at three heart valve centres: the Second Affiliated Hospital of Shantou University Medical College, the First Affiliated Hospital of Sun Yat-sen University, and the Affiliated Hospital of Guangdong Medical University. The participants were allocated to high-risk or low-risk groups using a cut-off value of 4% of EUROSCORE II. AVR Surgery, aortic valve replacement surgery. AS, aortic valve stenosis.

The study was conducted according to the principles of the Declaration of Helsinki and was approved by the Ethics Review Committee of the Shantou University Medical College. All the data were collected from the hospital electronic medical records, and follow-up was conducted through hospital visits or phone calls. Verbal informed consent was obtained from the participants, as approved by the Ethics Review Committee of the Second Affiliated Hospital of Shantou University Medical College(ERB number: ethics form 9-5).

## Data collection and definitions

Participants self-reported age, sex, height, body mass, smoking and alcohol history, and medication upon admission. History of hypertension, diabetes, coronary heart disease (CHD), atrial fibrillation, extra-cardiac artery lesions, or stroke was further validated by medical professionals on the basis of blood test or imaging results. Subsequent AVR surgery information was obtained from electronic medical records or telephone interviews. BMI was calculated as body mass (kg)/height (m)2.

Hypertension was defined by antihypertensive drug use, systolic blood pressure ≥140 mmHg, or diastolic blood pressure ≥90 mmHg. Chronic kidney disease (CKD) was defined as estimated glomerular filtration rate (eGFR) <60 mL/min/1.73 m2 using CKD-EPI equation [20].

Laboratory tests within 24 hours of admission included white blood cell count, haemoglobin, creatinine, uric acid, total cholesterol, low-density lipoprotein-cholesterol (LDL-C), high-density lipoprotein-cholesterol (HDL-C), amino-terminal brain-natriuretic peptide precursor (NT-proBNP), and troponin. The use of the following drugs was recorded: anti-platelet drugs, statins, angiotensin-converting enzyme inhibitors (ACEIs), angiotensin receptor blockers (ARBs), β-blockers, calcium channel blockers (CCBs), hypoglycaemic drugs, oral anticoagulants, and diuretics. The following echocardiographic indices were also recorded: left ventricular ejection fraction (LVEF), the maximum velocity of blood passing through the aortic valve (AV_Vmax), the mean gradient across the aortic valve (AV_MG), the aortic valve area (AVA), pulmonary arterial hypertension (PAH), aortic regurgitation, mitral regurgitation, and mitral stenosis.

## Definitions of echocardiographic indices

Transthoracic echocardiography followed American Society of Echocardiography guidelines[21]. AS severity was defined as [3,4]: (1) moderate AS: 1 cm2<AVA≤1.5 cm2, 3 m/s<Vmax≤ 3.9 m/s, or <20 mm Hg<MG≤39 mm Hg; and (2) severe AS: AVA≤1 cm2, Vmax≥4 m/s, or MG≥40 mm Hg. PAH was defined as a pulmonary arterial pressure ≥25 mm Hg, according to the Guidelines for the Diagnosis and Treatment of Pulmonary Hypertension in China (2021 edition)[22].

## Outcomes

The primary endpoint of the study was all-cause mortality from the time of the diagnosis, whose information was collected by trained medical staff through telephone contact with the participants or their families. The secondary endpoints were cardiovascular-related mortality and aortic stenosis-related mortality. The duration of follow-up was date of AS diagnosis to date of death or end of follow-up **Statistical analysis**

Continuous datasets were tested for normality using the Shapiro–Wilk method, and the data are presented as as mean ±SD for normal distribution, or median and interquartile range for skewed data. Categorical data are presented as counts and percentages. Homogeneous datasets compared using ANOVA, heterogeneous using Kruskal–Wallis test. P ≤ 0.05 was considered statistically significant. The participants were allocated to high-risk or low-risk groups using a cut-off value of EUROSCORE II of 4%, according to the 2017 ESC/EACTS guidelines[23]. Student’s t-test or ANOVA was used to evaluate differences between groups with respect to continuous data, and Pearson’s chi-square test or Fisher’s exact test was used to compare categorical datasets, as appropriate.

Kaplan–Meier method calculated cumulative survival, evaluated with log-rank test. Cox proportional hazard analyses were performed, generating hazard ratios (HRs) and 95% confidence intervals (CIs) for the relationships for EUROSCORE II and AVR status association with all-cause mortality. Next, four multivariable models were involved adjustment for potential confounders of all-cause mortality. Model 1 was adjusted for smoking status, alcohol consumption status, diabetes, atrial fibrillation, hypertension, gout, systolic blood pressure, and body mass. Model 2 was adjusted for the parameters in model 1, with the addition of WBC, RBC, PLT, LDLC, TG, TBIL, NTproBNP, troponin, and albumin. Model 3 was adjusted for the parameters in model 2, plus aortic valve deformity, rheumatic heart disease, AV-Vmax, AV-MG, the degree of AVA mitral insufficiency, and mitral stenosis. Model 4 was adjusted for the parameters in Model 3, with the addition of anti-platelet agent, statin, ACEI, ARB, β-blocker, CCB, diuretic, insulin, oral antidiabetic drug, and oral anticoagulant use. Further subgroup analyses were performed after the stratification of the participants according to baseline sex, age (≤ 70 or >70 years), BMI (<24 or ≥24 kg/m^2^), diabetes, hypertension, CHD, and the severity of AS (moderate or severe), to assess the consistency of the prognostic use of EUROSCORE II for all-cause mortality. Finally, the ROC curve was also used to determine the optimal cut-off value of EUROSCORE II for the prediction of all-cause mortality in the participants. All analyses were conducted in the IBM SPSS 26.0 statistical software and a two-sided P value<0.05 was considered statistically signifcant.

## Results

### Baseline characteristics of the participants

#### Clinical characteristics of the participants, stratified according to EUROSCORE II and AVR status

A total of 1,033 patients who met the diagnostic criteria for AS were initially included in the study, but 70 were excluded because they fulfilled one of the exclusion criteria, 219 because of missing EUROSCORE II data, and 200 because of missing covariant data. Thus ultimately, data for 544 participants were analysed in the present study (Fig. 1), comprising 300 (55.2%) men and 244 (44.9%) women, with a mean age of 64.7±13.0 years. The baseline characteristics of the participants, after being categorized according to the recommended cut-off value (4%) of EUROSCORE II and whether or not AVR was performed, are shown in Table 1.

**Table 1.**
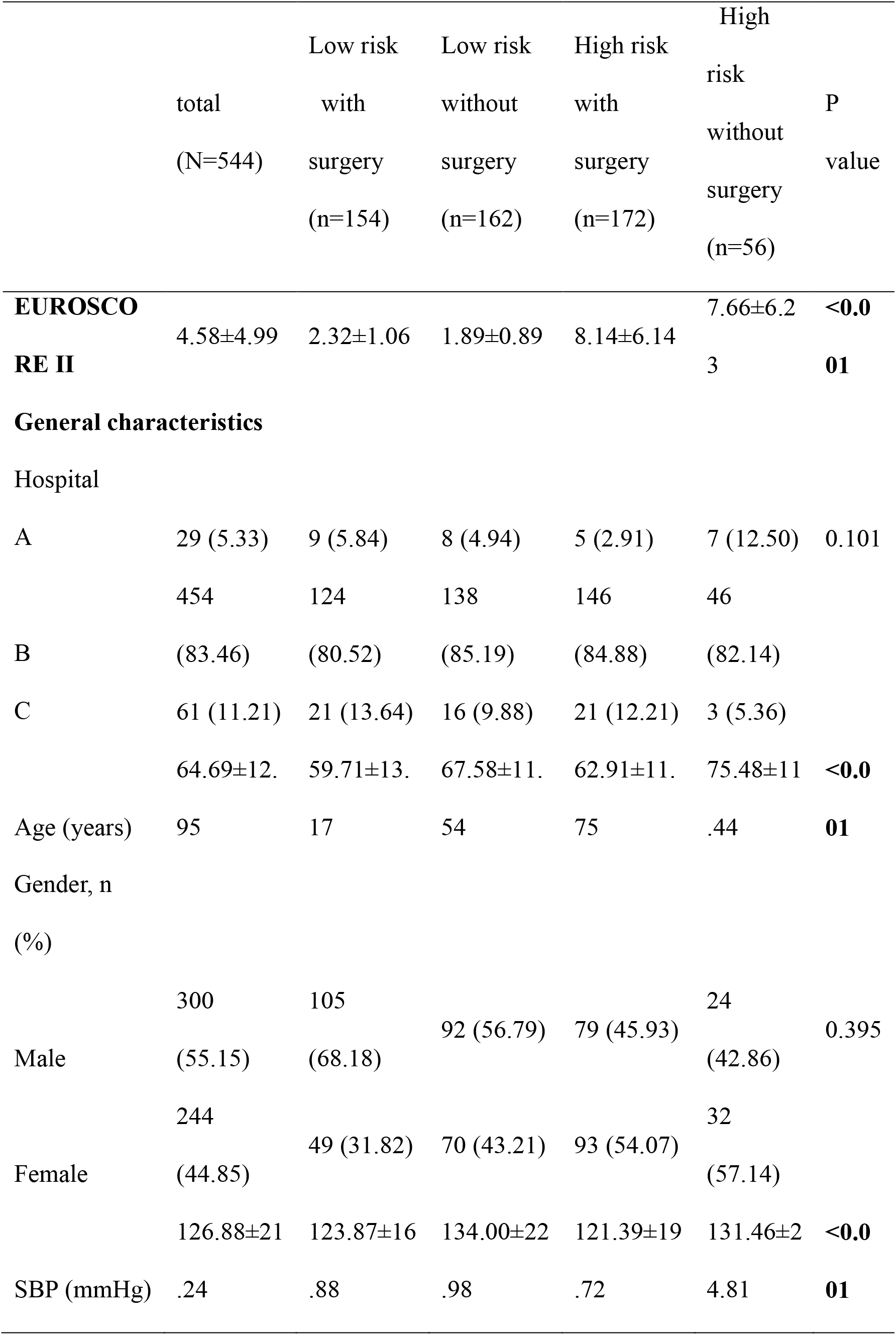
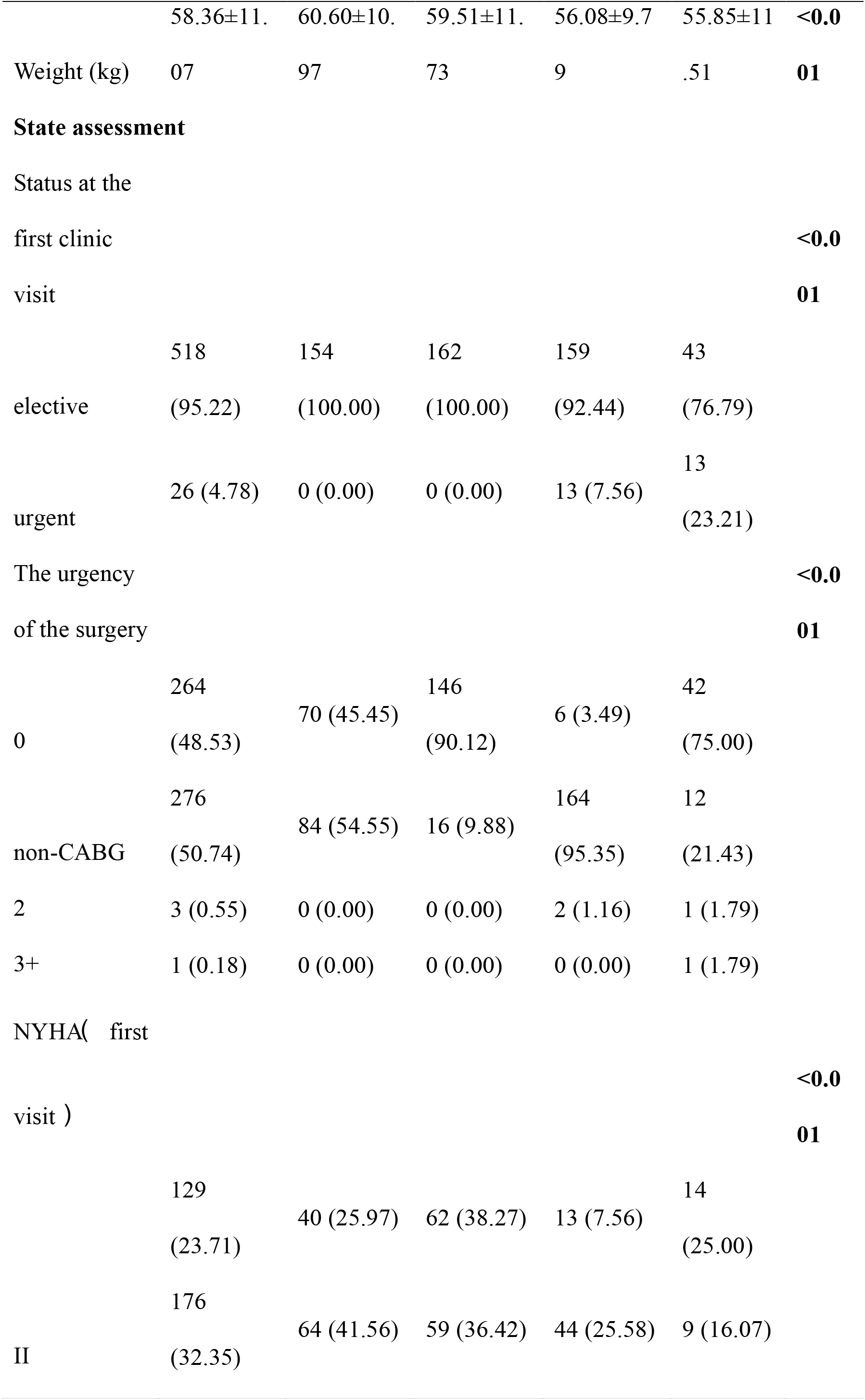
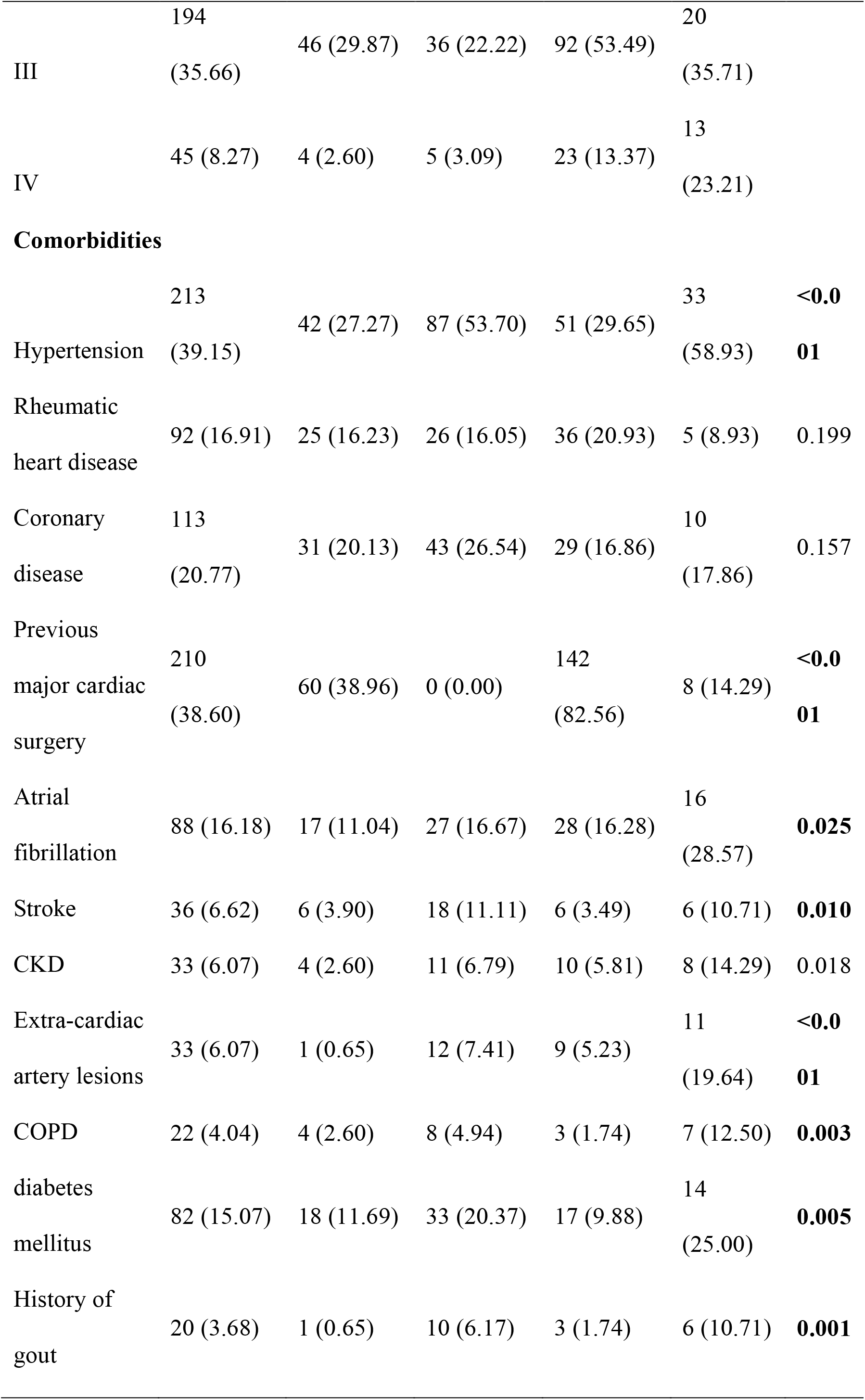
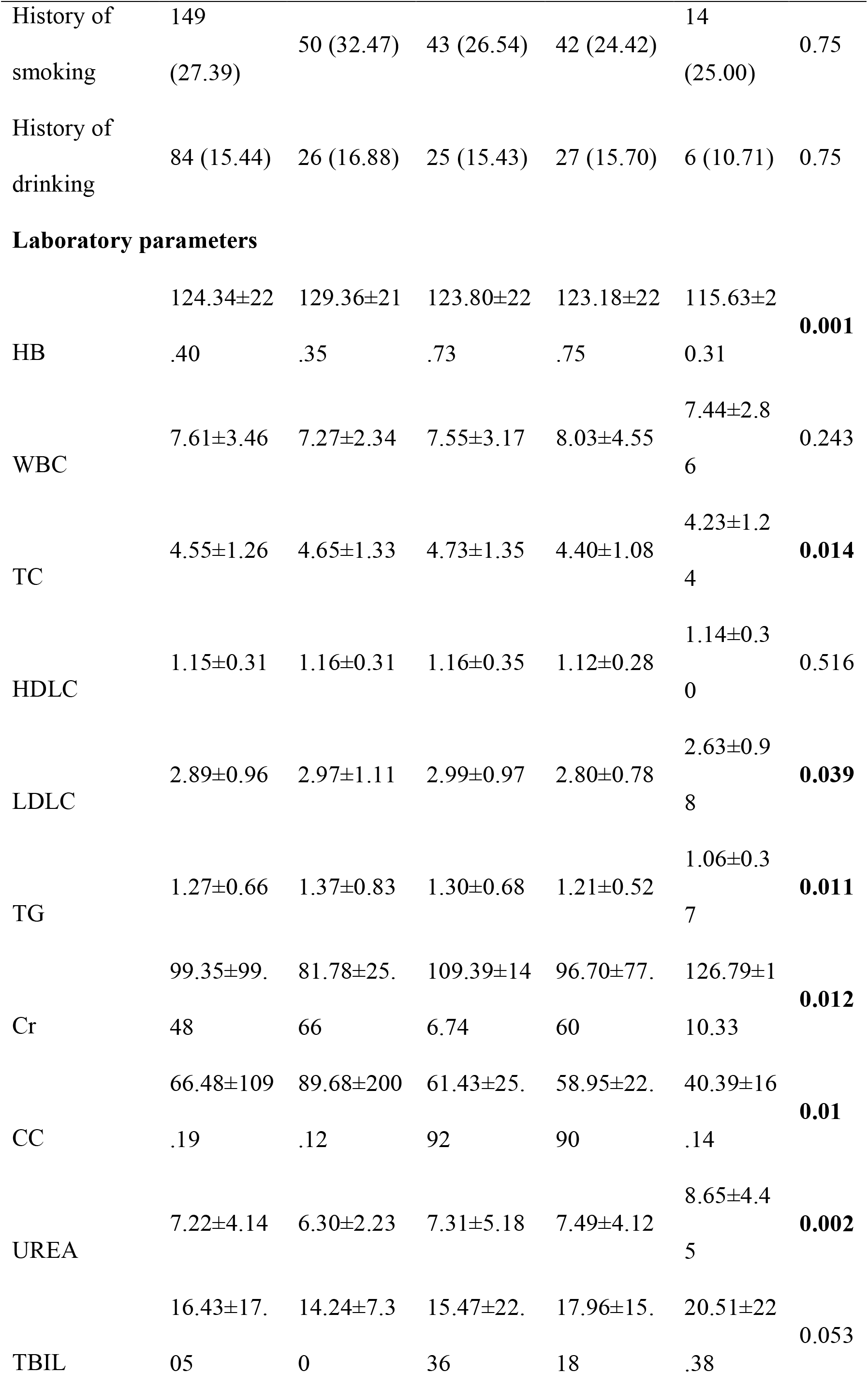
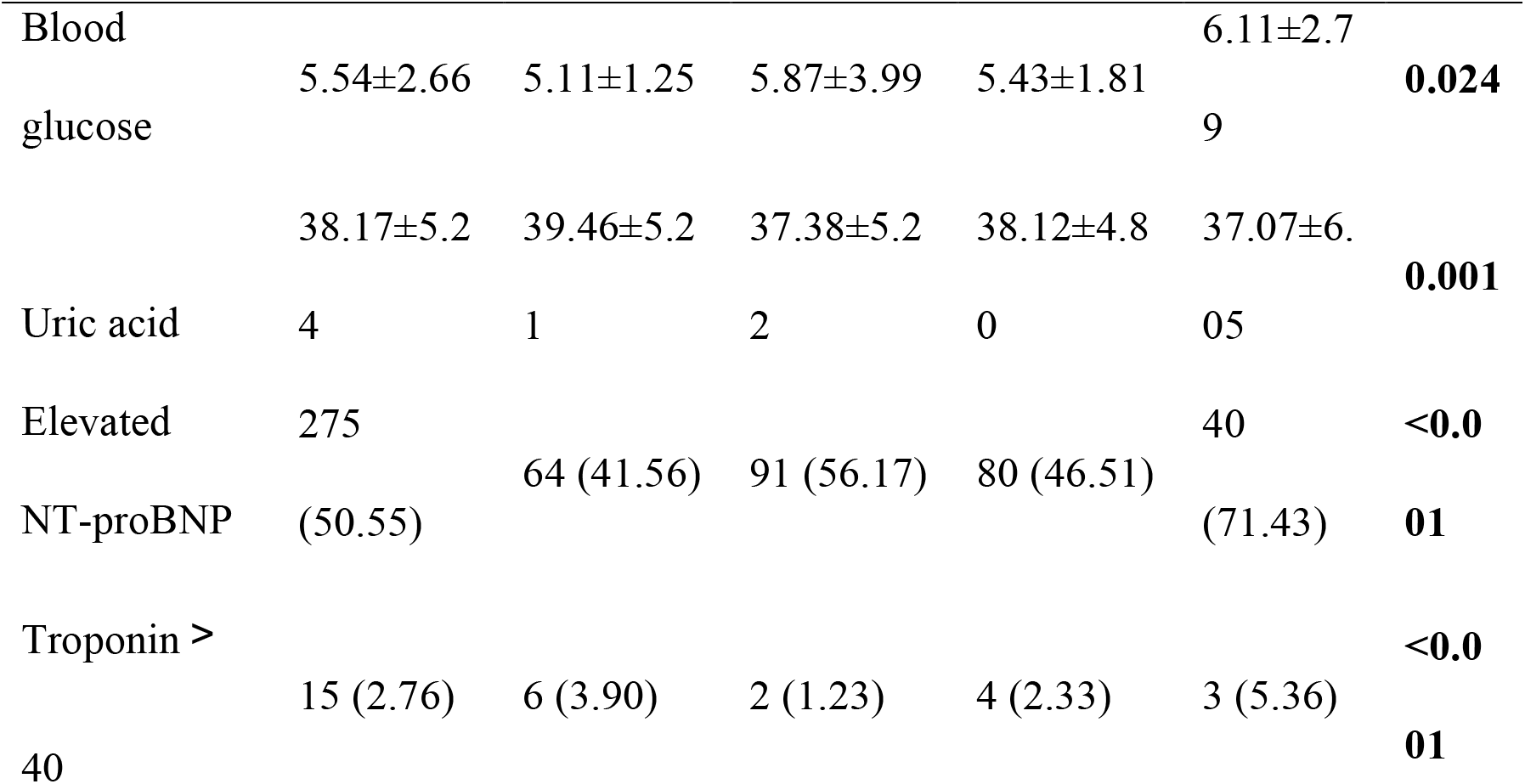
Clinical characteristics of the participants, stratified according to their EUROSCORE II and their AVR status

We evaluated the following six types of characteristics of the participants in the four groups: their general characteristics, preoperative status assessment, comorbidities, laboratory parameters, echocardiographic findings, and medical therapy. Those with a EUROSCORE II index ≥4% who did not undergo AVR surgery tended to be older, have higher serum Cr, TBIL, and blood glucose concentrations, to have a high NYHA score, and to have more comorbidities, such as diabetes and hypertension, but to have a lower body mass and LDL-C and TG concentrations than those in the other groups (all *P* ≤ 0.05). In addition, the echocardiographic findings for the participants (Additional file Table S1) showed that the participants in this group were more likely to have other valvular diseases, severe PAH, and lower LVEF. In addition, these participants were more likely to be administering statins, insulin, and oral anticoagulants. However, there were no differences among the four groups with respect to sex, their history of alcohol consumption or smoking, or in their oral anti-diabetic drug use (all *P*>0.05).

### Baseline characteristics of the participants, stratified according to only EUROSCORE II

During a follow-up period of 10.7 years, 132 (21.5%) participants had reached an endpoint. To evaluate the utility of EUROSCORE II alone for the prediction of all-cause mortality in patients with moderate-to-severe AS, we compared two groups of participants, categorized according to the recommended cut-off value (4%). As shown in Additional file Table S2, participants with higher EUROSCORE II (≥4%) were more likely to be female, older, require surgery more urgently, have more serious cardiac dysfunction and were more likely to have other heart valve diseases, such as mitral stenosis, mitral regurgitation, and aortic regurgitation. However, there were no significant differences between the groups with respect to the incidences of comorbidities (*P*>0.05).

## Relationships of all-cause mortality with EUROSCORE II and AVR status

### Results of the Kaplan–Meier analysis of the participants, stratified according to both EUROSCORE II and AVR status

The log-rank test was employed to compare the cumulative survival of the groups of participants, as depicted in Fig. 2. The analysis demonstrated that the participants in the group with high EUROSCORE II and who had not undergone AVR were at the highest risk of all-cause mortality of the four groups (log-rank *P*<0.001).

**Fig. 2.**
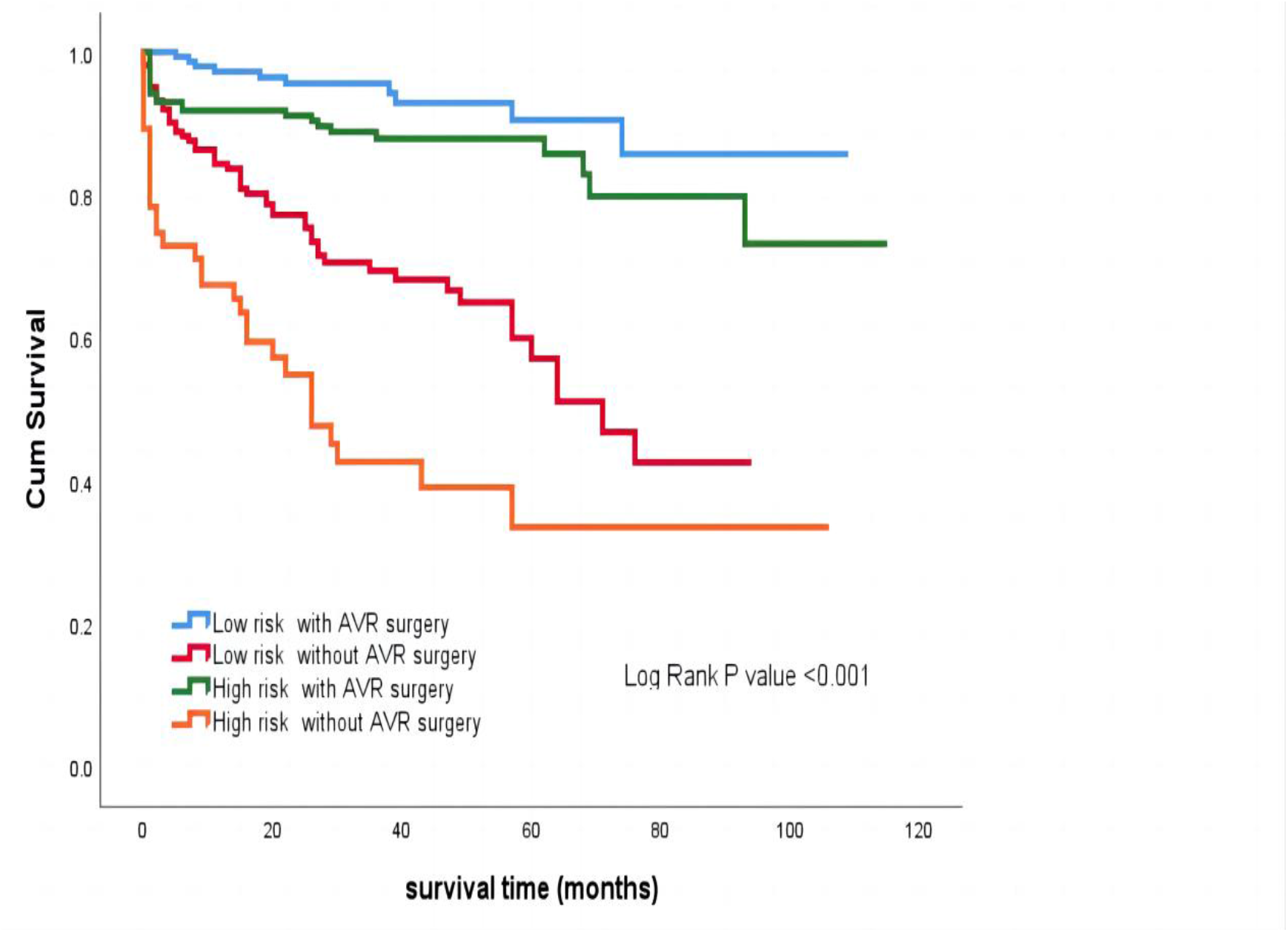
Kaplan–Meier survival curves for the participants, categorized according to their EUROSCORE II and their AVR status

### Results of the Kaplan–Meier analysis of the participants who had/hadn’t undergone AVR, stratified according to their EUROSCORE II

Compared with the low-risk group, the participants in the high-risk group showed significantly poorer cumulative survival, according to the Kaplan–Meier curves (*P*=0.001) (Additional File Fig. S1) . Similar results were obtained for the participants who had not undergone AVR (*P*=0.001) (Additional File Fig. S2).

### Comparison of the Kaplan–Meier curves for participants who did or did not undergo AVR

For the low-risk group of participants (EUROSCORE II <4%), the Kaplan–Meier curves and the log-rank test showed that the cumulative survival time significantly differed between those who had or had not undergone AVR (*P*=0.01) (Additional File Fig. S3). Similarly, among high-risk participants (EUROSCORE II ≥4%), those who had not undergone AVR (P<0.01) (Additional File Fig. S4) showed significantly poorer cumulative survival time than participants undergone AVR.

However, there was no difference between those who underwent SAVR or TAVI (*P*=0.692) (Additional File Fig. S5).

## Predictors of all-cause mortality

The findings of the univariate and multivariate Cox regression analyses of the relationships of EUROSCORE II and AVR status with the all-cause mortality of the participants are presented in Table 2. Univariate regression analysis showed that age, CAD, CKD, COPD, diabetes, atrial fibrillation, hypertension, albumin, critical status, urgency of surgery, EF, pulmonary hypertension, statin use, ACEI/ARB use, and insulin use were predictors of all-cause mortality.

**Table 2.**
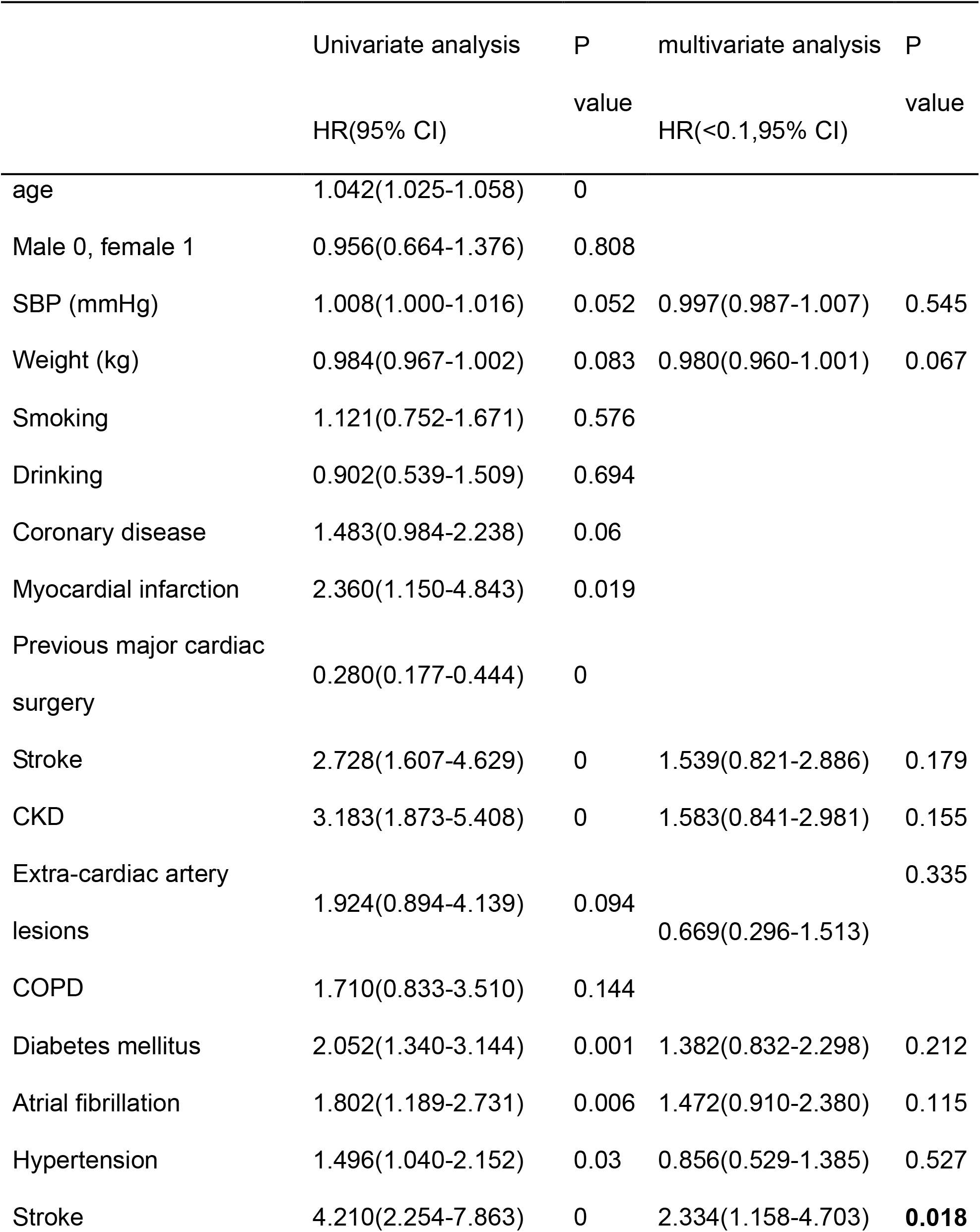
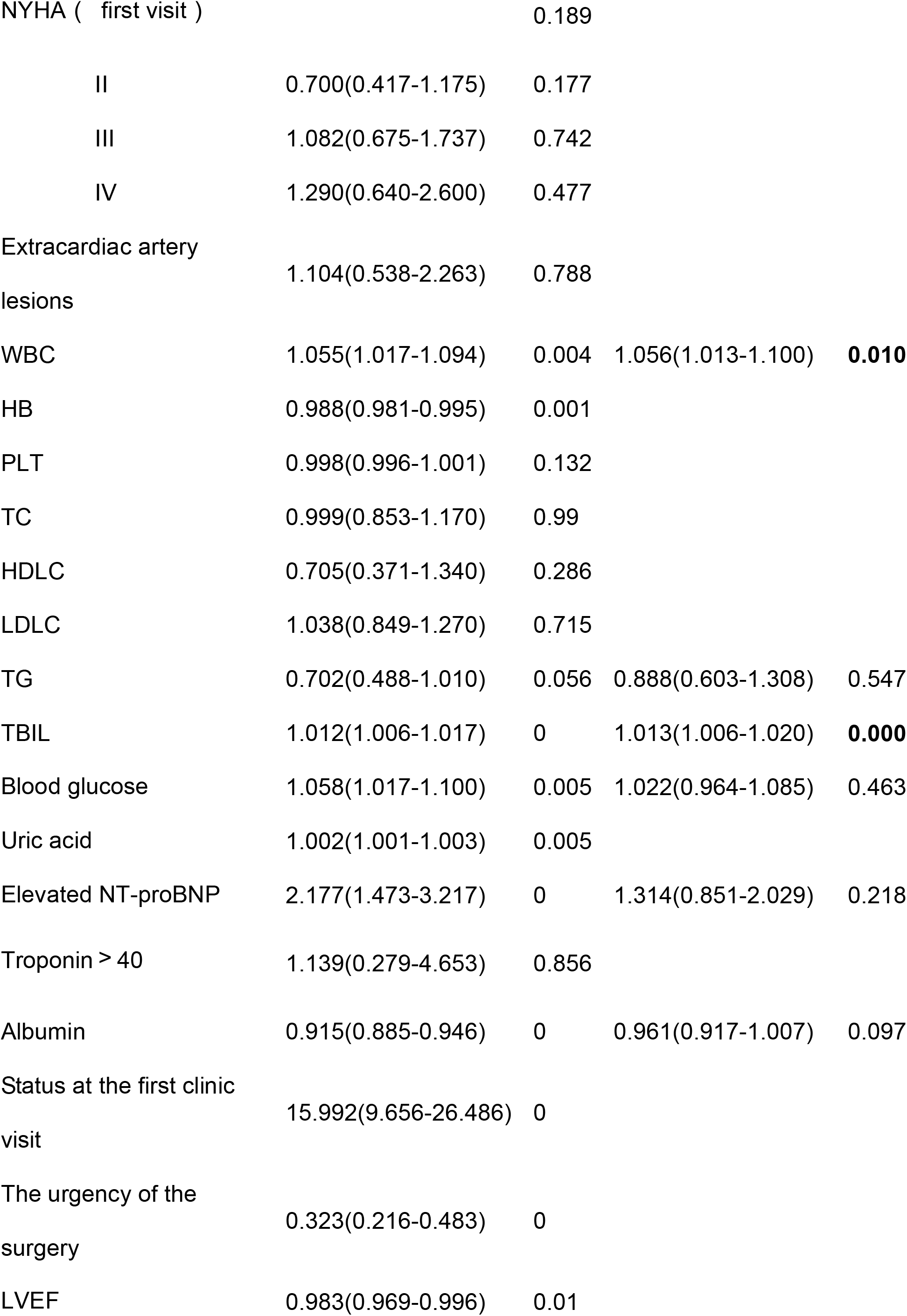
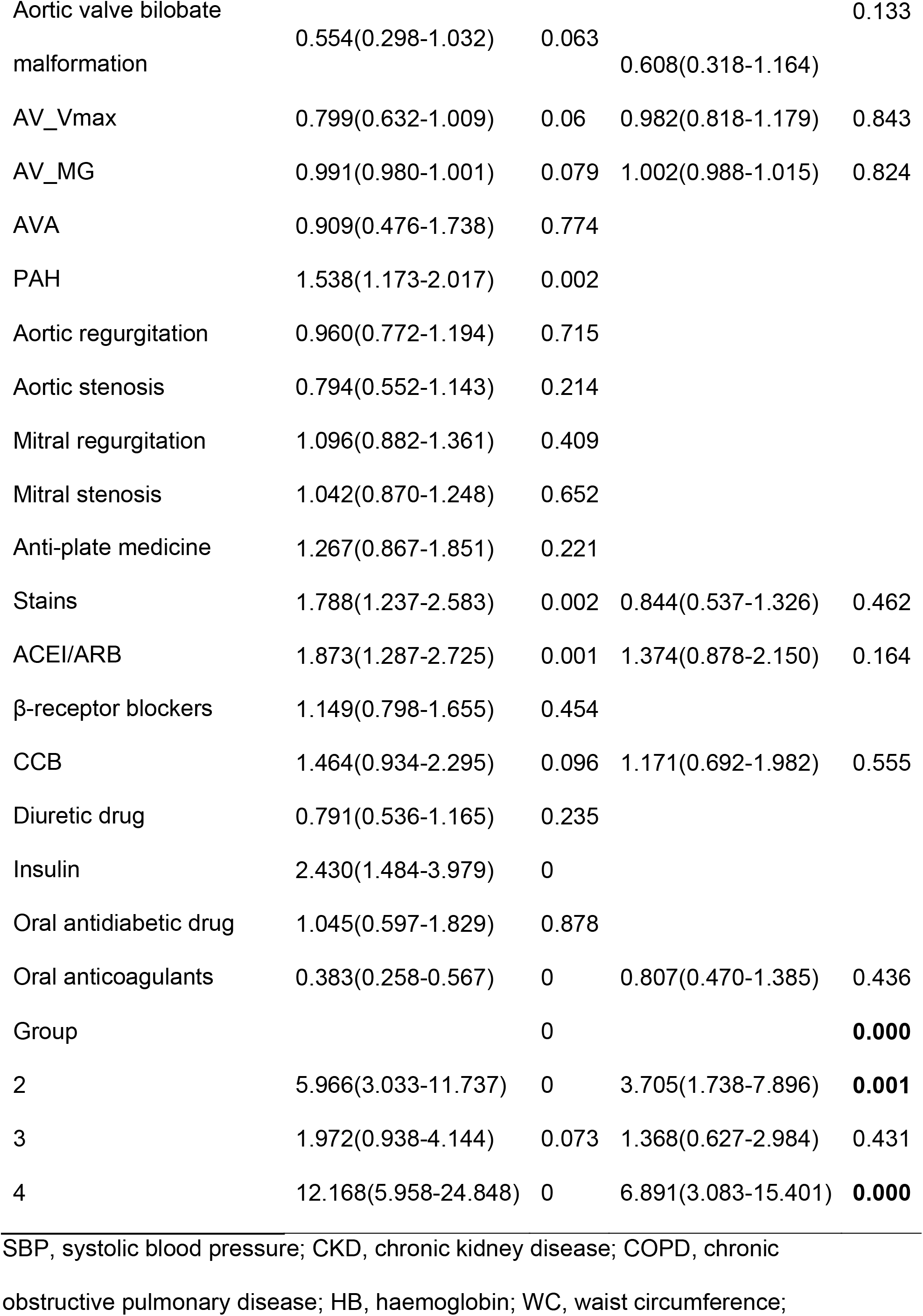
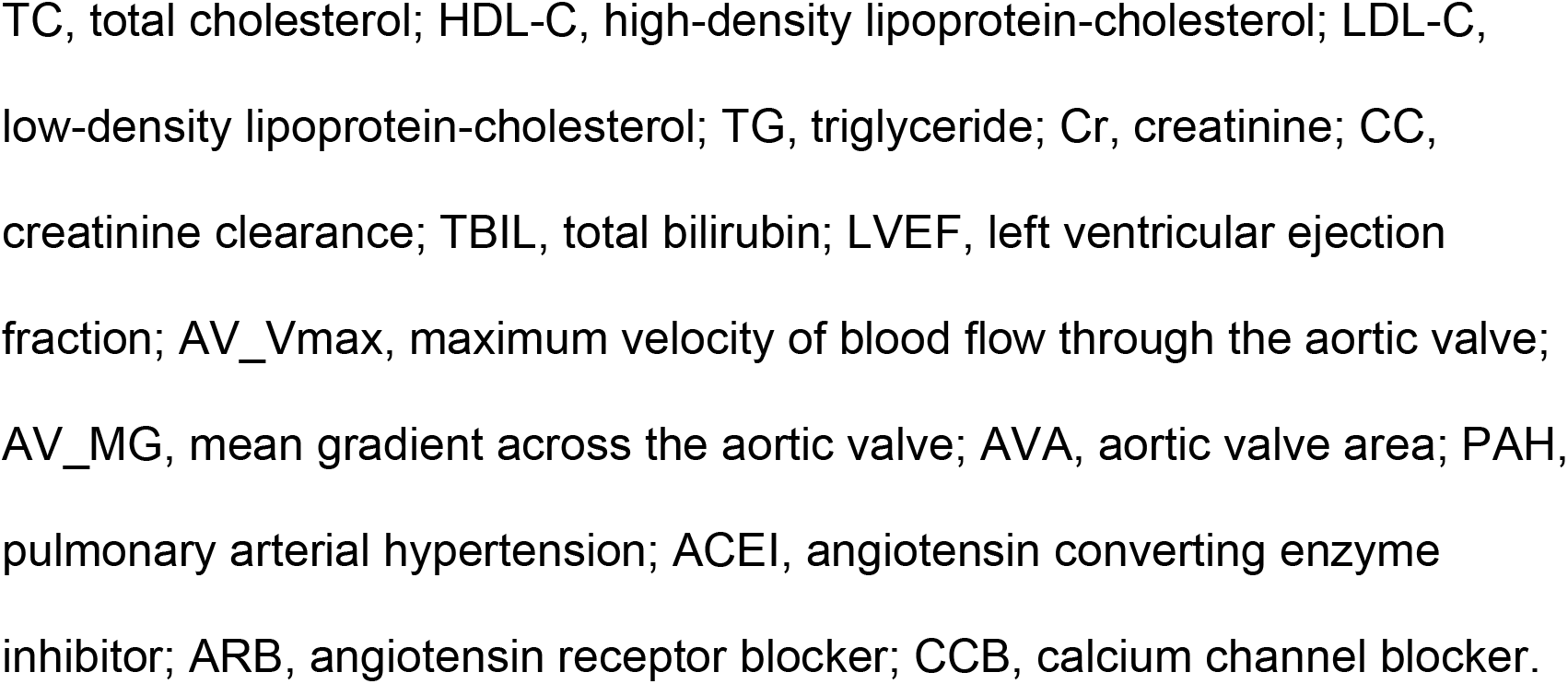
Results of the univariate and multivariate Cox analyses of the associations with EUROSCORE II and AVR status

The univariate analysis showed that group 2 (EUROSCORE II <4%, without AVR) were associated with a 5.97-fold higher risk than group 1 (EUROSCORE II <4%, with AVR) (HR, 5.966; 95% CI 3.033–11.737; *P*<0.001). Furthermore, participants in group 4 (EUROSCORE II ≥4% and no AVR) were at a 12.17-fold higher risk of all-cause mortality *vs*. those in group 1 (HR, 12.168; 95% CI 5.958–24.848; *P*<0.001).

Multivariate Cox regression analyses showed that group 1 (EUROSCORE II <4%, had undergone AVR) exhibited the lowest risk (HR, 3.705, 95% CI 1.738–7.896, *P*=0.001; and HR, 6.891, 95% CI 3.083–15.401, *P*<0.001 groups 2,4, respectively compared with group 1).

However, there no significant difference between participants in group 3 (EUROSCORE II ≥4% and AVR) and group 1 (HR, 1.368; 95% CI 0.627–2.984, *P*=0.431). Notably, significant and more steady results were obtained after further adjustment for potential confounders in Models 1–4 (Table 3). The fully adjusted HR and 95% CI in Model 4 for groups 2–4 *vs*. group 1 were 3.492 (1.583–7.703), 1.333 (0.605–2.936), and 6.605 (2.817–15.486), respectively (*P*<0.05).

**Table 3.**
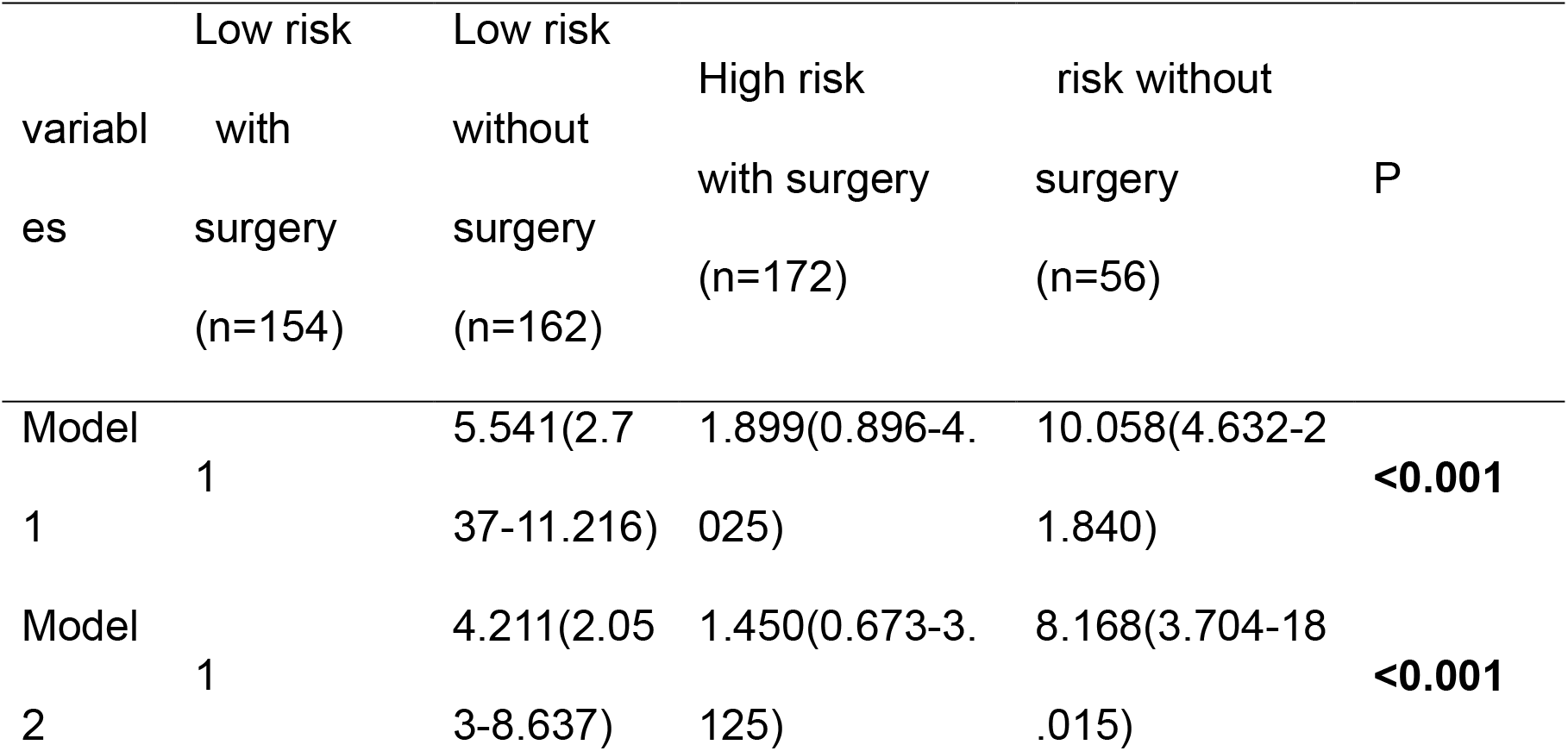
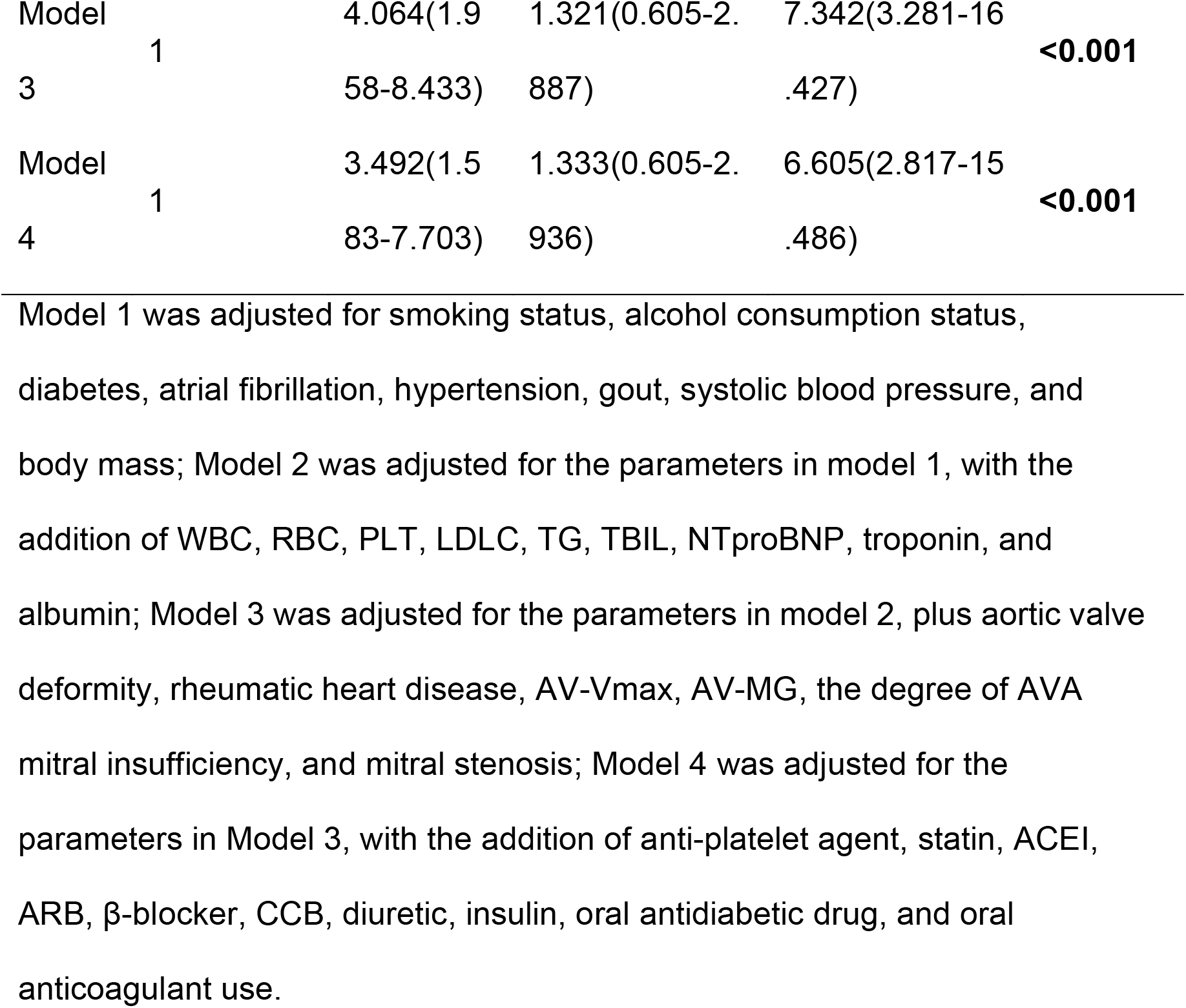
Relationships of EUROSCORE II and AVR status with the all-cause mortality of the participants

Moreover, participants in high risk without surgery (group 4) were further adjusted as the reference to prove that, among high-risk participants (EUROSCORE II ≥4%), perform AVR (group 3) were associated with a 0.023-fold risk than without AVR (group 4) (HR, 0.023; 95% CI 0.118-0.857, *P*<0.01).

### Results of the subgroup analyses

To further validate the identified difference in risk between participants who had or had not undergone AVR, two subgroup analyses were performed to evaluate the effects of relevant parameters on the relationship between EUROSCORE II and the all-cause mortality of the participants. The participants were stratified according to sex (male or female), age (≤70 or >70 years), diabetes (present or absent), hypertension (present or absent), CHD (present or absent), severity of AS (moderate or severe), aortic regurgitation (none, mild, moderate, or severe), LVEF (<40%, 40%−50%, or >50%), and the use of hypoglycaemic medication (yes or no).

#### Results of the subgroup analysis of the participants who had not undergone AVR

The results showed stronger associations between EUROSCORE II and the risk of all-cause mortality in younger (≤70 years old) male participants who did not have diabetes, hypertension, or CHD, were not taking hypoglycaemic medication, had mild aortic regurgitation, had an LVEF <40%, and had moderate aortic stenosis (Fig. 3).

**Fig. 3.**
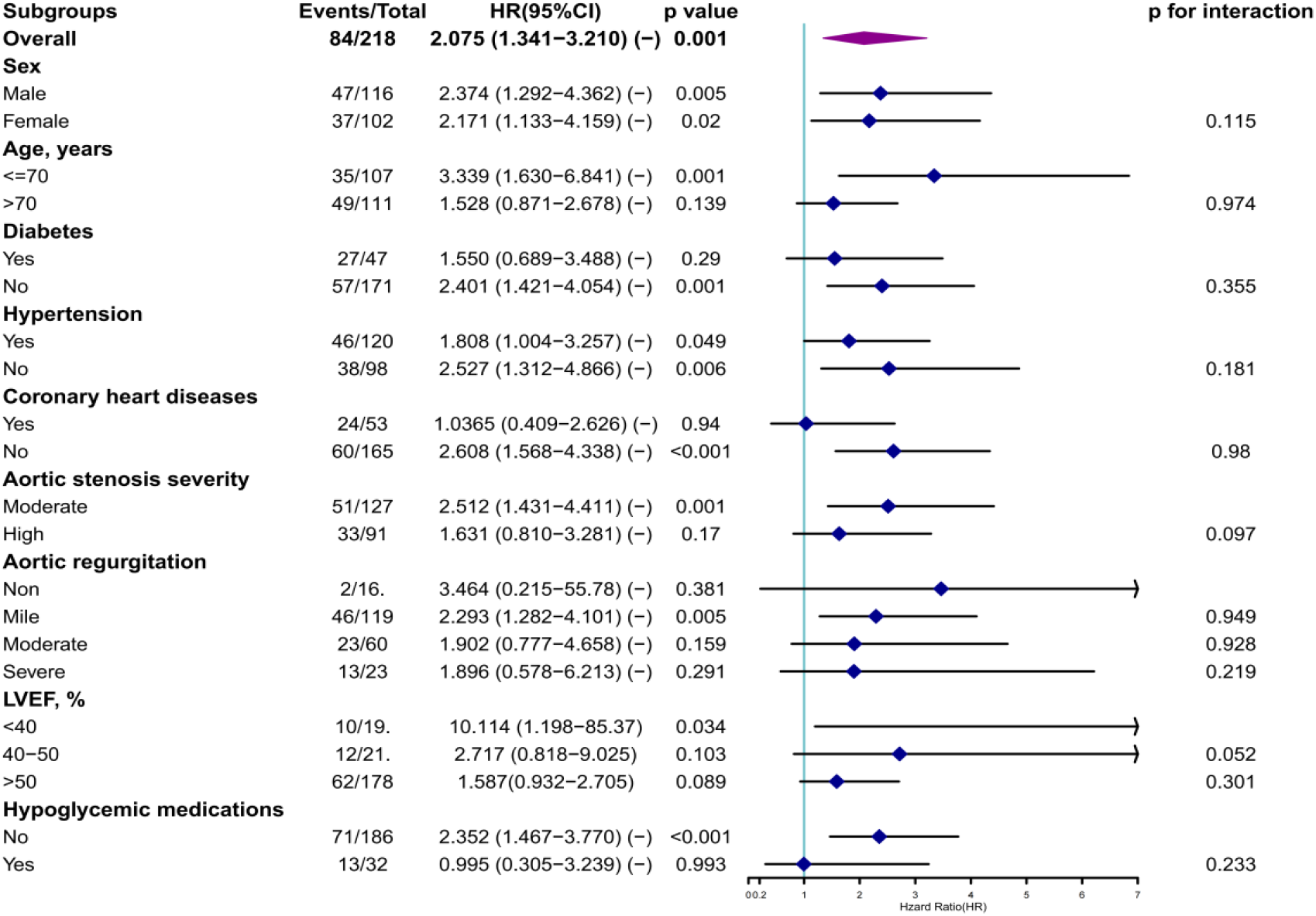
Results of the subgroup analysis of the association between EUROSCORE II and the lack of AVR HR, hazard ratio; CI, confidence interval.

The relationship of the EUROSCORE II index with all-cause mortality remained after adjustment for the confounders across all the subgroups, but there were no significant interactions between the subgroups (*P* for all interactions>0.05).

#### Results of the subgroup analysis of the participants who had undergone AVR

Analysis of the data for the participants who had undergone AVR (Additional file Fig. S6) revealed a similar association between EUROSCORE II and the risk of all-cause mortality risk in participants who did not have diabetes or CHD and were not taking hypoglycaemic medication. None of the subgroups showed significant interactions (*P* for all interactions>0.05).

### Optimal cut-off value of EUROSCORE II for the prediction of outcomes in the participants who had not undergone AVR

Fig. 4 shows the ROC curve analysis of the prognostic value of EUROSCORE II for the participants. The optimal cut-off value of EUROSCORE II identified for the prediction of the primary outcome in the participants who had not undergone AVR was 2.23%, which was associated with an AUC of 0.675 (95% CI: 0.609–0.74;

**Fig. 4.**
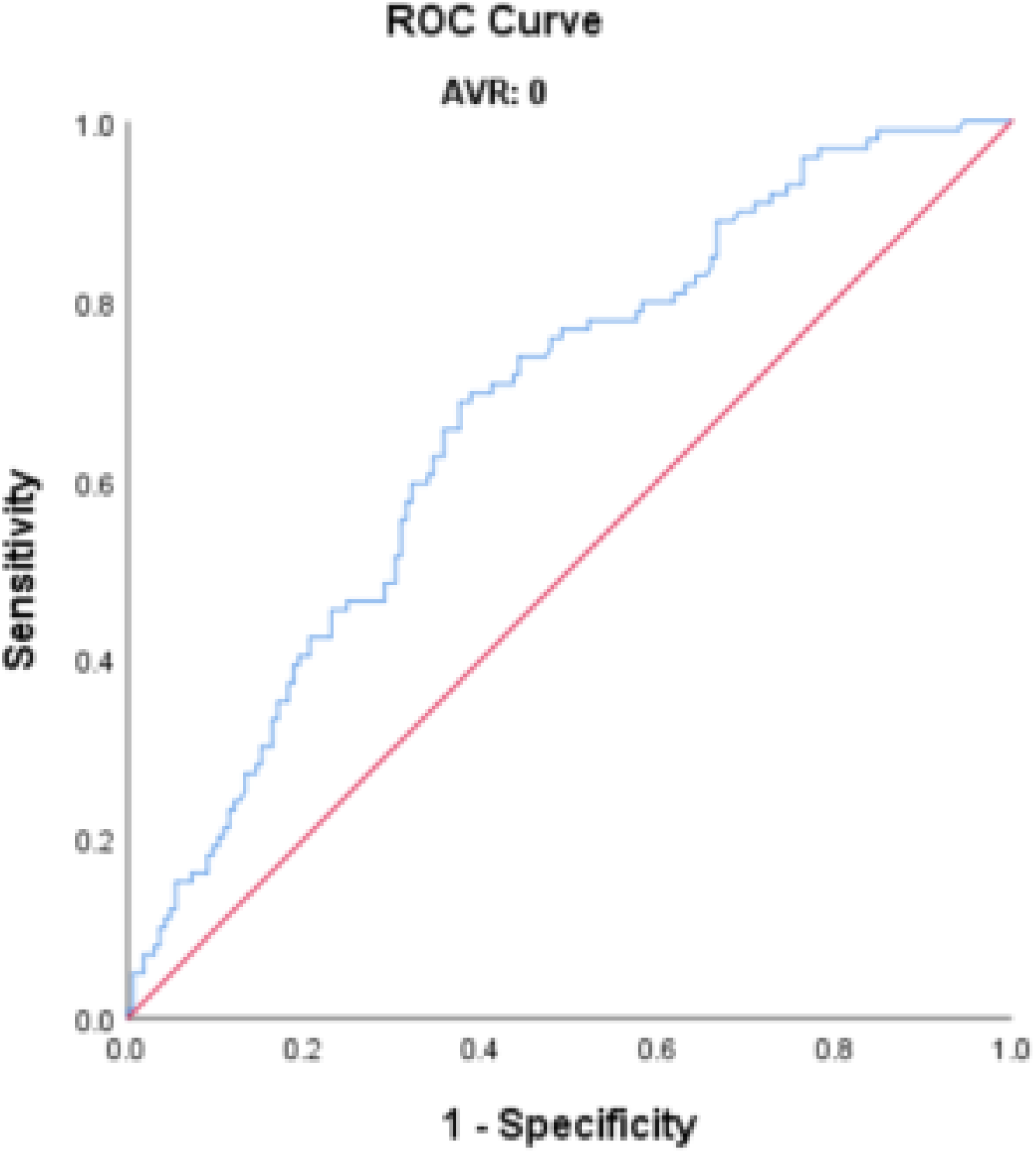
ROC curve for the use of EUROSCORE II to evaluate the prognosis of the participants who had not undergone AVR

*P*<0.001). Therefore, the predictive value of a EUROSCORE II threshold of 2.23% appears to be superior to that of 4% for the prediction of outcomes in patients without AVR.

We also performed Kaplan–Meier analysis and Cox multivariate analysis to confirm this finding. Kaplan–Meier analysis in supplemental Fig. S7 demonstrated that participants with a EUROSCORE II >2.23% had a higher risk of all-cause mortality than those with values <2.23% (log-rank, P<0.001). Additionally, compared with the lower EUROSCORE II group, the higher EUROSCORE II (≥2.23%) group had a 2.111-fold higher risk of all-cause mortality (HR 2.111, 95% CI (1.069–4.166), *P*=0.031), according to Cox multivariate analysis (Additional file Table S2).

## Discussion

A growing body of evidence demonstrates that patients with symptomatic severe aortic stenosis have a poor prognosis if they do not undergo surgery, with a mean survival time of <4 years[8,24]. Therefore, AVR is widely recommended for the treatment of such patients and is effective[24].

In the present study, we first explored the relationship between EUROSCORE II and the long-term prognosis of patients with moderate-to-severe AS. We found that a high EUROSCORE II is significantly associated with a higher risk of all-cause-mortality in such patients and AVR could reduce death risk to that of patients with a lower EUROSCORE II (<4%).

We first evaluated the long-term prognosis of patients who had received an initial diagnosis of moderate-to-severe AS within the preceding 10 years, and found that those with a EUROSCORE II ≥4%, whether or not they have undergone AVR, have a significantly higher risk of all-cause mortality than those with a EUROSCORE II <4%. Many previous clinical studies have shown that the EUROSCORE II has potential for use as a predictor of all-cause mortality for patients who undergo TAVI. A retrospective study of 350 patients with at least moderate AS who underwent TAVI showed that patients who died within 30 days had higher EUROSCORE II than those survived (12.6±1.8 *vs*. 7.5±0.3%, *P*<0.001). This was associated with an AUC of 0.70, which implies that EUROSCORE II is a good predictor of short-term mortality[25].

Another study of data obtained from 59 patients with symptomatic severe aortic stenosis who had undergone TAVI between 2010 and 2014 showed that EUROSCORE II is a predictor of in-hospital mortality and 30-day mortality[26]. However, the duration of follow-up in these studies was too short for the findings to be highly relevant to clinical practice. However, a recent study by Fan *et al*. classified 332 patients with low-gradient severe aortic stenosis and the preservation of LVEF into high-risk (EUROSCORE II ≥4%, n=115) and low-risk (EUROSCORE II <4%, n=208) groups and followed them over a 2-year period. They found that the cumulative survival of the high-risk group was significantly poorer than that of the low-risk group[27], which is consistent with the present findings that the use of 4% as a cut-off value has some predictive value over a period of >2 years.

Kaplan–Meier analysis showed that patients who undergo SAVR or TAVI to treat moderate-to-severe AS have comparable risks of relevant outcomes (*P*=0.692). This finding is in general consistent with the results of some recent studies. One prospective study of medium-term follow-up data from the Nordic Aortic Valve Intervention (NOTION) trial, which was obtained for 280 patients at three centres in Denmark and Sweden, showed that there was no difference in the all-cause mortality associated with TAVR or SAVR after 2 years (8.0% and 9.8%, respectively; *P*=0.54) or with respect to cardiovascular mortality (6.5% and 9.1%; *P*=0.40)[28]. In addition, another study of data obtained during the NOTION trial also showed no significant difference in the risk of all-cause mortality after 8 years of follow-up of these patients[29].

In the present study, which involved a duration of follow-up of up to 10 years, we also found evidence that patients with a low EUROSCORE II (<4%) should still be recommended to undergo SAVR or TAVR (*P*=0.001) to have a good prognosis, especially when there is some urgency in their condition. According to the 2021 ESC/EACTS guidelines for the management of valvular heart disease, SAVR is recommended for patients <75 years old, while TAVR is recommended for older patients (>75 years), high-risk patients (STS-PROM/EUROSCORE II >8%), and those who are not eligible for surgery (class I)[9]. Despite this, many previous randomized clinical trials have demonstrated that TAVR is non-inferior or even superior to SAVR with respect to both short- and medium-term outcomes[30-37].

Notably, the PARTNER2 study[40], the SURTAVI study[41], several studies of the use of TAVR in low-risk patients performed by the American College of Cardiology (ACC), including the PARTNER 3 study (using the SAPIEN-3 ultra transcatheter heart valve) and the Evolut Low Risk Trial study (using the self-expanding Evolut R valve)[42] showed that TAVR was associated with better than, or at least non-inferior, results to, SAVR in low-risk patients. After the 5-year follow-up period, the Deep Dive into the PARTNER 3 Five-Year Study showed that patients who undergo TAVR or SAVR have similar annual incidences of cardiovascular-related mortality, stroke, and re-hospitalization owing to cardiovascular disease (approximately 1%, 1%, and 3%, respectively). These findings are consistent with our conclusion that patients with low EUROSCORE II (<4%) should still be recommended to undergo SAVR or TAVR. Furthermore, some other studies have shown that SAVR and TAVR are safe and effective, and are associated with a low risk of severe aortic stenosis, a low risk of surgical complications, a short hospital stay, and a low risk of mortality during the perioperative and 30-day postoperative periods[38-39].

Finally, the most appropriate cut-off value of EUROSCORE II to be used prognostically varies according to the underlying disease, owing to the diversity in the risk factors and pathophysiological mechanisms associated[43-44]. However, several previous studies have shown that the use of EUROSCORE II is associated with an underestimation of the risk of mortality for high-risk patients and an overestimation of this risk for those with low levels of risk following various types of cardiac surgery[43-49]. A previous study of 12,325 patients who underwent major cardiac surgery showed that EUROSCORE II was a fairly accurate predictor up to a risk of 30%, but over-predicted outcomes above this level[50]. Furthermore, EUROSCORE II was shown to be a fairly accurate predictor for octogenarians up to a risk level of 10%, but overestimated the risks above this level[49].

Although EUROSCORE II has been shown to be a good predictor of in-hospital mortality following various types of surgery, the optimal cut-off values of EUROSCORE II for the prediction of postoperative mortality were 10%–20%[51]. The present ROC curve analysis for EUROSCORE II showed that the most suitable cut-off value for the prediction of outcomes in patients with moderate-to-severe AS who had not undergone AVR was 2.23% (AUC, 0.675), rather than 4%. Therefore, we next allocated the participants to a high EUROSCORE II group and a low EUROSCORE II group using this cut-off value and compared their risks of outcomes. Kaplan–Meier curve (log-rank *P*<0.001) and Cox multivariate (HR 2.111, 95% CI (1.069–4.166), *P*=0.031) analyses were performed to provide further evidence that 2.23% is the optimal value to differentiate these groups of patients. It is worth mentioning that the original value of 4% was used for patients with AS, whether or not they had undergone surgery, whereas the 2.23% value calculated in the present study is specific for patients with moderate-to-severe AS who have not undergone AVR.

The major strength of the present study is that it was a multi-centre observational study, the external validity of which could be verified. Furthermore, we followed the participants for up to 10 years, which renders the results more reliable. We found a significant association between EUROSCORE II and all-cause mortality in patients with moderate-to-severe AS after comprehensive adjustment for potential confounders, and made consistent findings in subgroup analyses. These findings should contribute to the existing body of knowledge regarding the clinical utility of EUROSCORE II to predict the outcomes of patients with AS, thereby leading to better outcomes and a lower future burden of this valvular disease.

## Limitations

The present study also had several limitations. First, the prediction of risk depends on the accurate evaluation of each risk factor in EUROSCORE II, but the scores used are based on European patients, rather than other groups. Therefore, the results need further validation in studies of patients of other ethnicities. Second, although the number of patients enrolled was relatively large and it was a multi-centre study, the numbers were relatively low in comparison to the those accessible from a large database, meaning that the analysis was relatively underpowered. Third, the echocardiographic and laboratory parameters were only assessed once, at admission, which may have resulted in bias owing to measurement errors or differences in criteria between the centres. Finally, according to the 2021 ESC/EACTS guidelines, SAVR should be recommended for younger patients with a low surgical risk (<75 years, STSPROM/EUROSCORE II <4%), while TAVR is recommended for older patients (>75 years) and those at high risk (STS-PROM/EUROSCORE II >8%), but we did not find a difference in the prognosis of the participants who underwent SAVR or TAVI (*P*>0.05), which may be attributable to the limited sample size.

## Conclusions

We have shown for the first time that both EUROSCORE II and whether or not AVR is performed are independently associated with the long-term prognosis of patients with moderate-to-severe AS. Moreover, those who receive an initial diagnosis of moderate-to-severe AS are recommended to undergo surgery (SAVR or TAVR) early in their disease course, regardless of their EUROSCORE II. Finally, we have identified an optimal EUROSCORE II cut-off value of 2.23% for the determination of the prognosis of patients with moderate-to-severe AS who have not undergone AVR.

## Data Availability

The datasets used and/or analyzed during the current study are available from the corresponding authors upon reasonable request.

## Ethical approval

This study was approved by the Ethics Review Committee of the Second Affiliated Hospital of Shantou University Medical College(ERB number: ethics form 9-5).

Patient follow-up was conducted via telephone contact, with verbal informed consent approved by the institutional ethics committee.

## Pre-registered clinical trial number

Pre-registered clinical trial number: NCT06069232.

## Consent for publication

Not applicable.

## Acknowledgments

We thank Mark Cleasby, PhD from Liwen Bianji (Edanz) (www.liwenbianji.cn) for editing the language of a draft of this manuscript.

## Central Message

Our heart valvular disease intervention center construction unit from The Second Affiliated Hospital of Shantou University Medical College is part of the National Clinical Medical Research Center for Radiology and Therapy, Interventional Center for Valvular Disease, Dongfang Huaxia Cardiovascular Health Institute, Suzhou Industrial Park.

Our data were from Three heart valve centers, which are derived from a database called Aortic Valve Diseases RISk facTOr assessmenT and Prognosis modeL Construction (ARISTOTLE).

## Glossary of Abbreviations

EuroSCORE II (European System for Cardiac Operative Risk Evaluation), SBP (systolic blood pressure), CKD (chronic kidney disease), COPD (chronic obstructive pulmonary disease), HB (haemoglobin), TC (total cholesterol), HDL-C (high-density lipoprotein-cholesterol), LDL-C(low-density lipoprotein-cholesterol), TG (triglyceride), Cr (creatinine), CC(creatinine clearance), TBIL(total bilirubin), LVEF (left ventricular ejection fraction), AV_Vmax(maximum velocity of blood flow through the aortic valve), AV_MG (mean gradient across the aortic valve), AVA (aortic valve area), PAH(pulmonary arterial hypertension), ACEI (angiotensin converting enzyme inhibitor), ARB(angiotensin receptor blocker), CCB(calcium channel blocker), LVEF (left ventricular ejection fraction),AV_Vmax(maximum flow velocity through the aortic valve), AV_MG(mean gradient across the aortic valve), AVA(aortic valve area), PAH(pulmonary arterial hypertension)

